# Polygyny among men in Ghana: a population based cross-sectional study

**DOI:** 10.1101/2024.11.19.24317603

**Authors:** Gladys Quartey, Abigail Padi, Louis Kobina Dadzie, King David Dzirasah, Richard Gyan Aboagye, Bright Opoku Ahinkorah, Abdul-Aziz Seidu

**Author notes:** **Corresponding author** KDD. **Email addresses** GQ AP LKD RGA BOA AS.

## Abstract

**Background:** Polygyny has been practiced in many cultures around the world over the past couple of decades. Available evidence indicates that polygyny is practiced in more than 83% of the 849 cultures globally. Therefore, this study seeks to assess the prevalence and determinants of polygyny among men in Ghana.

**Methods:** This research is based on 1,892 married men using the 2014 Ghana Demographic and Health Survey (GDHS) data. Polygyny was the outcome variable. Multilevel mixed-effects logistic regression models were fitted to examine the factors associated with polygyny. The results were presented as adjusted odds ratio with 95% confidence interval.

**Results:** The prevalence of polygyny was 8.8[7.5%-10.5%]. Men aged 45-59[aOR=7.01; 95%CI= [2.87,17.15] and 30-44 [aOR=4.36; 95%CI= [1.81,10.53] were more likely to engage in polygyny compared to those aged 15-29. Men in communities with high wealth status had higher odds of polygyny compared to those in communities with low wealth status [aOR=2.49; 95%CI= [1.22,5.09]. On the other hand, men with a secondary level of education [aOR=0.53;95%CI= [0.29,0.95] and those with comprehensive knowledge of HIV and AIDS [aOR=0.59;95%CI= [0.37,0.94] were less likely to engage in polygyny compared to those with no formal education and those without comprehensive HIV and AIDS knowledge.

**Conclusion:** The study has contributed to knowledge on the prevalence and determinants of polygyny among men in Ghana. In Ghana, age, education, community wealth status, and knowledge of comprehensive HIV/AIDS are determinants of polygyny among men. Interventions to improve universal access to education and social protection policies are critical to reshaping socio-cultural practices and views that engender polygyny among men in Ghana. In addition, strengthening existing laws and policies is important in addressing the social challenges caused by polygamous unions. Furthermore, explorative studies must be conducted to identify the possible impact of polygamy on those who practice it.

## Background

Polygyny is a form of marriage that involves a husband having multiple wives concurrently and is the most common form of polygamous marriage [1]. Polygyny has been practiced in many cultures around the world over the past couple of decades. Available evidence indicates that polygyny is practiced in more than 83%of the 849 cultures globally [2,3, 4]. In numerous low- and middle-income countries, polygyny is a distinguishing feature of family life. Between 20% and 40% of women in various West African countries, including Nigeria, Côte d’Ivoire, Cameroon, Mali, and Benin claim to be in polygynous marriages [5].

Polygyny in Africa has been linked to a lack of men in post-slave trade Africa, while others argue that polygyny has been a cultural norm in Africa for centuries and predates the advent of colonists [5, 6]. Formal marriage laws and the predominant religion of the Christian colonists, Christianity, seem to have reduced polygyny to a more cultural role [7]. Polygyny continues to exist on the continent, even though figures on polygynous marriages indicate that the number of polygynous marriages is decreasing and this could be attributed to its harmful consequences on the health of women; hence, the urgent call for its abolishment [8, 5]. With polygyny, there are certain advantages, such as the sharing of farm or home labour and a rise in fertility, particularly in countries with high infant death rates [9, 10, 11]

Possible factors that may be associated with polygyny vary from economic, such as wealth disparities between men and women, to demographic, such as skewed sex ratios resulting from greater male death rates owing to risky vocations, to political, such as war and revolution [6]. A study done in [12] Ethiopia found that age, religion, educational background, and residence are some of the factors associated with polygyny among men. For instance, no education or lower level of education was positively associated with polygyny as compared to those with a higher level of education. When it comes to age also, older men were more likely to find themselves in polygynous relations as compared to those who are younger. In relation to religion, being a Muslim was positively associated with polygyny [12].

In Ghana, a man may marry one woman by Ordinance and as many as he wishes by customary marriage, as long as the culture permits it [13, 14, 15). Ghana is situated within what is known as the “polygyny belt” which stretches from West Africa to East Africa [16]. The prevalence of polygyny within the “polygyny belt” is generally high [1]. The high prevalence of polygyny in sub-Sahara Africa is a cause of worry as the phenomenon has been associated with some health and social problems [18, 19]. Firstly, it has been observed that polygyny contributes to the transmission of HIV/AIDS due to unprotected extramarital coitus [20]. Secondly, polygyny has been identified to significantly contribute to the incidence of child mortality. Scholars have argued that children within polygynous unions or marriages tend to have nutritional and health challenges due to under-nutrition and unavailability of financial resources [16]. Thirdly, polygyny has also been identified to be associated with intimate partner violence. Partners in polygynous unions are subjected to emotional and physical abuse [1].

Despite these, within the Ghanaian context, there is a knowledge gap on the extent of polygyny among men and the possible factors for it. Determining the prevalence and the factors associated with polygyny among men in Ghana is critical in the realization of Sustainable development Goals (SDGs) 3 and 5 that seek to ensure health and well-being for all, at every stage of life and achieve equality among all genders and empower women and girls by 2030, respectively [21]. Despite the social, public health, and developmental significance of polygyny in Ghana, there is less concentration on polygyny among men in Ghana. Therefore, this study seeks to assess the prevalence and determinants of polygyny among men in Ghana.

## Methods

### Data Source

The men’s recode file of the 2014 Ghana Demographic and Health (GDHS) dataset was used in this study. The GDHS is a nationally representative survey whose major goal is to generate accurate data on topics such as socioeconomic factors, fertility, and family planning. The survey is cross-sectional and employs a two-staged sample design, with the first stage entailing the selection of clusters made up of enumeration areas (EAs) defined in the Population and Housing Census (PHC). A systematic sample of houses is done in the second step. The Ghana Statistical Service (GSS) and Ghana Health Service (GHS) conducted the study with funding from the US Agency for International Development (USAID). Detailed decription of the sampling and methodology can be found in the published report [22].

### Study setting and study population

Ghana is located in the Centre of West Africa’s coast, with a land area of 238,537 square kilometers. It is bordered on the east by Togo, on the north by Burkina Faso, on the west by Cote d’Ivoire, and on the south by the Gulf of Guinea. Moreover half of Ghana’s population lives in the Greater Accra and Ashanti administrative regions. The 2014 GDHS included 4,388 men, however, for this study only 1,892 married men who had complete information on all the variables of interest were considered for study.

### Variables

#### Outcome variable

The study’s outcome variable “Polygyny” was derived from the question “how many wives/partners do you have?” This varied from 0 to 4, but was divided into two categories: “0-1=0” and “2 or more=1 “ [12].

#### Explanatory variables

The independent variables were grouped into individual and community factors. The individual factors included the age of respondents, educational level, wealth quintile, religion, age at first sex, and comprehensive HIV knowledge. Community-level factors included residence, region, community educational level, and community poverty. Age was recoded as “15-29”, “30-44”and “45-59”. Religion was recoded into Christianity, Islam, traditionalist, and no religion. Age at first sex was also grouped as below 18 years and above 18 years. Comprehensive knowledge of HIV was composed of a set of questions on knowledge of HIV transmission and misconception. Using the characteristics of the males in the clusters, the community-level determinants of education and poverty were aggregated.

#### Data processing and analyses

The data analysis procedure began with data cleaning and recategorization of variables for better comprehension. Frequencies and percentages were used to offer descriptive analyses of the various variables of importance. Bivariate analyses were also used to determine the relationships between the different independent factors and the dependent variable. To assess the normality of the community-level components produced, we used the Shapiro Wilk test of normality and the histogram. We utilized a multilevel mixed-effect logistic regression analysis to assess the individual and community level characteristics related to polygyny based on the hierarchical nature of the GDHS. In total, four models were utilized. We started with a model with no covariates (model 0), then added the individual variables (model 1). Model 3 contained community-level variables, whereas Model 4 is the final model, which incorporates all variables, including individual and community-level variables. We used the survey command to compensate for over and under-sampling in all of our analyses. The threshold for statistical significance was established at p<0.05. We used Stata 14 for all analyses.

## Results

### Background characteristics of respondents

Table 1 shows the background characteristics of the respondents. About 53% were aged 30-44, 55.5% had secondary level of education, 26.6% were in the richest wealth quintile. It was also shown that 67.6% are Christians and 50.6% are in rural areas. It was also found that polygyny was highest among those aged 45-49 (11.5%), those with no education(25.1%), those in the poorest wealth quintile (25.8%), traditionalist(28.4%), those in rural areas (10.7%) and those in Northern Region (30%). The prevalence of polygyny was 8.8%[7.5%-10.5%] among men in Ghana.

**Table 1.** Background characteristics of respondents and prevalence of polygyny.

|  |  |  |  |  |
| --- | --- | --- | --- | --- |
| **Variable** | **Freq (%)** | **Polygyny (8.8% [CI=7.5-10.4%])** |  | **P-value** |
|  |  | No | Yes |  |
|  |  | % (95%CI) | %(95%CI) |  |
| **Age** |  |  |  | < 0.001 |
| 15-29 | 197(10.4) | 96.8 [93.3-98.5] | 3.2 [1.5-6.7] |  |
| 30-44 | 1,000(52.9) | 91.7 [89.7-93.4] | 8.3 [6.6-10.3] |  |
| 45-59 | 695(36.7) | 88.5 [85.8-90.8] | 11.5 [9.2-14.2] |  |
| **Educational level** |  |  |  | < 0.001 |
| No formal education | 357(18.9) | 74.9 [70.0-79.3] | 25.1 [20.7-30.0] |  |
| Primary | 227(12) | 89.9 [85.5-93.1] | 10.1 [6.9-14.5] |  |
| Secondary | 1,051(55.5) | 95.2 [93.6-96.4] | 4.8 [3.6-6.4] |  |
| Higher | 257(13.6) | 97.5 [93.9-99.0] | 2.5 [1.0-6.1] |  |
| **Wealth index** |  |  |  | < 0.001 |
| Poorest | 381(20.1) | 74.2 [69.9-78.2] | 25.8 [21.8-30.1] |  |
| Poorer | 343(18.1) | 91.2 [87.8-93.7] | 8.8 [6.3-12.2] |  |
| Middle | 310(16.4) | 94.1 [91.0-96.2] | 5.9 [3.8-9.0] |  |
| Richer | 356(18.8) | 96.8 [93.8-98.4] | 3.2 [1.6-6.2] |  |
| Richest | 502(26.6) | 97.8 [95.5-99.0] | 2.2 [1.0-4.5] |  |
| **Religion** |  |  |  | < 0.001 |
| Christianity | 1,280(67.6) | 95.6 [94.2-96.7] | 4.4 [3.3-5.8] |  |
| Islam | 398(21) | 82.9 [78.5-86.6] | 17.1 [13.4-21.5] |  |
| Traditionalist | 113(6) | 71.6 [64.3-77.9] | 28.4 [22.1-35.7] |  |
| No religion | 101(5.3) | 88.1 [77.8-94.0] | 11.9 [6.0-22.2] |  |
| **Age at first sex** |  |  |  | 0.889 |
| <18 | 463(24.5) | 91.2 [88.0-93.7] | 8.8 [6.3-12.0] |  |
| >=18 | 1,429(75.5) | 91 [89.3-92.5] | 9 [7.5-10.7] |  |
| **Comprehensive knowledge on HIV** |  |  |  | < 0.001 |
| No | 1,258(66.5) | 89.2 [87.2-90.9] | 10.8 [9.1-12.8] |  |
| Yes | 634(33.5) | 94.9 [92.8-96.4] | 5.1 [3.6-7.2] |  |
| **Type of place of residence** |  |  |  | < 0.001 |
| Urban | 929(49.1) | 95.3 [93.2-96.8] | 4.7 [3.2-6.8] |  |
| Rural | 963(50.9) | 87 [84.4-89.3] | 13 [10.7-15.6] |  |
| **Region** |  |  |  | < 0.001 |
| Western | 192(10.1) | 95.1 [91.1-97.3] | 4.9 [2.7-8.9] |  |
| Central | 193(10.2) | 96.9 [92.4-98.8] | 3.1 [1.2-7.6] |  |
| Greater accra | 373(19.7) | 96.1 [91.7-98.2] | 3.9 [1.8-8.3] |  |
| Volta | 153(8.1) | 85.2 [76.9-90.9] | 14.8 [9.1-23.1] |  |
| Eastern | 169(8.9) | 94.7 [89.6-97.4] | 5.3 [2.6-10.4] |  |
| Ashanti | 322(17) | 97.5 [94.5-98.8] | 2.5 [1.2-5.5] |  |
| Brong ahafo | 151(8) | 91 [85.5-94.5] | 9 [5.5-14.5] |  |
| Northern | 202(10.7) | 70 [61.3-77.5] | 30 [22.5-38.7] |  |
| Upper east | 87(4.6) | 80.1 [73.0-85.8] | 19.9 [14.2-27.0] |  |
| Upper west | 50(2.7) | 84.6 [73.7-91.5] | 15.4 [8.5-26.3] |  |
| **Community educational status** |  |  |  | < 0.001 |
| Low | 1,241(65.6) | 95.3 [93.5-96.6] | 4.7 [3.4-6.5] |  |
| High | 651(34.4) | 83.1 [79.8-85.9] | 16.9 [14.1-20.2] |  |
| **Community Poverty level** |  |  |  | < 0.001 |
| Low | 1,218(64.4) | 96.8 [95.3-97.9] | 3.2 [2.1-4.7] |  |
| High | 674(35.6) | 80.7 [76.4-84.4] | 19.3 [15.6-23.6] |  |

### Factors associated with polygyny among men in Ghana

Table 2 shows the results on the factors associated with polygyny among men in Ghana. Men aged 45-59 [aOR=7.01; 95%CI= [2.87,17.15] and 30-44 [aOR=4.36; 95%CI= [1.81,10.53] were more likely to engage in polygyny compare to those aged 15-29. Thosein communities with high wealth status had higher odds of polygyny compared to those in communities with low wealth status [aOR=2.487;95%CI= [1.22,5.09]. On the other hand, men with secondary level of education [aOR=0.53; 95%CI= [0.29,0.95] and those with comprehensive knowledge of HIV and AIDS [aOR=0.59; 95%CI= [0.37,0.94] were less likely to engage in polygyny compared to those with no formal education and those without comprehensive HIV and AIDS knowledge.

**Table 2:** Factors associated with polygyny among men in Ghana.

| Table 2: Factors associated with polygyny among men in Ghana |  |  |  |  |
| --- | --- | --- | --- | --- |
| Variable | Model 0 | Model 1<br>aOR[95%CI] | Model 2<br>aOR[95%CI] | Model 3<br>aOR[95%CI] |
| **Age** |  |  |  |  |
| 15-29 |  | 1 [1.00,1.00] |  | 1 [1.00,1.00] |
| 30-44 |  | 4.13** [1.71,9.98] |  | 4.36**<br>[1.81,10.53] |
| 45-59 |  | 5.85***<br>[2.40,14.26] |  | 7.01***<br>[2.87,17.15] |
| **Education** |  |  |  |  |
| No formal education |  | 1 [1.00,1.00] |  | 1 [1.00,1.00] |
| Primary |  | 0.52* [0.30,0.92] |  | 0.65 [0.35,1.19] |
| Secondary |  | 0.47** [0.28,0.77] |  | 0.53* [0.29,0.95] |
| Higher |  | 0.48 [0.18,1.29] |  | 0.42 [0.15,1.16] |
| **Wealth quintile** |  |  |  |  |
| Poorest |  | 7.32***<br>[3.24,16.52] |  | 2.74 [0.83,9.06] |
| Poorer |  | 3.13** [1.39,7.06] |  | 1.71 [0.55,5.30] |
| Middle |  | 2.33 [1.00,5.44] |  | 1.95 [0.70,5.37] |
| Richer |  | 1.19 [0.49,2.91] |  | 1.24 [0.49,3.13] |
| Richest |  | 1 [1.00,1.00] |  | 1 [1.00,1.00] |
| **Religion** |  |  |  |  |
| Christianity |  | 0.82 [0.38,1.76] |  | 0.77 [0.35,1.68] |
| Islam |  | 1.99 [0.91,4.33] |  | 1.71 [0.77,3.83] |
| Traditionalist |  | 1.75 [0.75,4.09] |  | 1.14 [0.48,2.71] |
| No religion |  | 1 [1.00,1.00] |  | 1 [1.00,1.00] |
| **Comprehensive HIV knowledge** |  |  |  |  |
| No |  | 1 [1.00,1.00] |  | 1 [1.00,1.00] |
| Yes |  | 0.66 [0.42,1.04] |  | 0.59* [0.37,0.94] |
| **Residence** |  |  |  |  |
| Urban |  |  | 1 [1.00,1.00] | 1 [1.00,1.00] |
| Rural |  |  | 1.24 [0.74,2.08] | 1.12 [0.61,2.04] |
| **Region** |  |  |  |  |
| Western |  |  | 1 [1.00,1.00] | 1 [1.00,1.00] |
| Central |  |  | 0.47 [0.15,1.49] | 0.55 [0.17,1.78] |
| Greater Accra |  |  | 1.22 [0.47,3.15] | 1.73 [0.61,4.90] |
| Volta |  |  | 2.12 [0.85,5.30] | 2.49 [0.94,6.56] |
| Eastern |  |  | 0.74 [0.26,2.13] | 0.72 [0.24,2.12] |
| Ashanti |  |  | 0.47 [0.17,1.32] | 0.51 [0.18,1.50] |
| Brong ahafo |  |  | 1.09 [0.41,2.85] | 1.05 [0.39,2.85] |
| Northern |  |  | 3.10* [1.25,7.68] | 2.51 [0.96,6.57] |
| Upper east |  |  | 1.58 [0.58,4.30] | 1.38 [0.47,4.00] |
| Upper west |  |  | 1.31 [0.41,4.15] | 1.19 [0.36,3.97] |
| **Community educational level** |  |  |  |  |
| Low |  |  | 1 [1.00,1.00] | 1 [1.00,1.00] |
| High |  |  | 1.33 [0.78,2.27] | 0.80 [0.43,1.49] |
| **Community wealth** |  |  |  |  |
| Low |  |  | 1 [1.00,1.00] | 1 [1.00,1.00] |
| High |  |  | 3.84***[2.08,7.08] | 2.487* [1.22,5.09] |
| **Random effect results** |  |  |  |  |
| PSU variance (95%CI) | 1.59(0.96, 2.64) | 0.46 (0.16, 1.32) | 0.29(0.08, 1.03) | 0.29(0.08,1.12) |
| ICC | 0.33 | 0.12 | 0.08 | 0.08 |
| Wald chi-square | Ref | 148.29*** | 124.38*** | 159.61*** |
| **Model fit** |  |  |  |  |
| Log-likelihood | -543.22 | -456.15 | -474.47 | -438.66 |
| AIC | 1090.45 | 942.3 | 976.9 | 931.3 |
| Exponentiated coefficients; 95% confidence intervals in brackets |  |  |  |  |
| * p<0.05, ** p<0.01, *** p<0.001 |  |  |  |  |

## Discussion

Using the 2014 Ghana Demographic and Health Survey data, this study examined the prevalence and determinants of polygyny among men in Ghana. We found a prevalence of 8.8% of polygyny among men in Ghana. The prevalence is much lower than in other sub-Saharan African and developing countries [23, 24, 25]. For instance, Amone and Arao (2014) found 35% prevalence of polygyny among men in Northern Uganda. Also, in a study conducted in Pakistan [25] found 51.8% prevalence of polygyny among men in Pakistan.

Age was significantly associated with increased odds of men engaging in polygyny in Ghana. Those age 45 to 59 had higher odds of engaging in polygyny than those age 15 to 29. This is congruent with findings from previous studies [25, 26]. A possible explanation for this is the effect of modernization in changing cultural norms that promote polygamous marriage within Ghanaian society [27]. This might influence the younger cohort to be less inclined to engage in polygamous unions compared to the older cohort [28]. Relatedly, education is significantly associated with higher odds of men engaging in polygyny in Ghana. Men with secondary education have lower odds of engaging in polygyny compared with men with no education. This is consistent with findings from previous studies done in Ethiopia [12] and Pakistan [25]. It may be that formal education is associated with westernization which predominantly promotes cultural practices that engender monogamous relationship norms [27]. In addition, it is also possible that the tendency of formal education on promoting small family size may influence those educated to prefer monogamous unions compared to those with no education [29].

In terms of comprehensive HIV Knowledge, men with no comprehensive HIV knowledge had higher odds of engaging in polygyny compared with men with comprehensive HIV knowledge. This finding does not deviate from existing literature [12]. This may be explained by the fact that those with comprehensive HIV knowledge are aware of the significance of multiple sexual partners in the spread of the virus [30]. Community wealth was significantly associated with higher odds of engaging in polygyny. This finding deviates from existing literature [12]. However, those in communities with high wealth quintile had higher odds of engaging in polygyny compared to those in communities with lower wealth quintile. A probable reason for this is the availability of communal resources to support multiple wives in wealthy communities [31, 32].

## Strength and Limitations

The study has strengths and limitations. In relation to strength, a large sample size was used for the study. This allows for the findings to be generalized. Also, the data collected is nationally representative. This is as a result of the multi-stage sampling procedure used. In addition, the data is reliable and this is a result of the utilization of experienced data collectors in the data collection process. In relation to limitations, due to the cross-sectional nature of the study, causality cannot be established. Also, there is the possibility of overestimation or underestimation due to self-reporting. Moreover, the Demographic and Health Survey Data used is old but it is the more current data collected in Ghana.

## Conclusion

The study has contributed to knowledge on the prevalence and determinants of polygyny among men in Ghana. In Ghana, age, education, community wealth status, and knowledge on comprehensive HIV/AIDS are determinants of polygyny among men. Interventions to improve universal access to education and social protection policies are critical to reshaping socio-cultural practices and views that engender polygyny among men in Ghana. In addition, strengthening existing laws and policies is important in addressing the social challenges caused by polygamous unions. Furthermore, explorative studies must be conducted to identify the possible impact of polygamy on those who practice it.

## Data Availability

Data available upon request

## Availability of data and materials

Dataset utilised and analysed for the study is available from the corresponding author subject to resequest.

## Acknowledgement

We acknowledge Ghana Statistical Service, Ghana Health Service and ICF International for making the data available to us.

## Contribution

All authors (GQ, AP, LKD, KDD, RGA, BOA and AS) contributed to the drafting and review of the manuscript.

## Corresponding author

King David Dzirasah

## Ethics declarations

Since the data was obtained from Ghana Statistical Service (GSS) and IFC, ethical clearance was granted by the respective institutional review boad of Ghana Statistical Service and IFC.

## Consent for publication

Not applicable

